# Exploring barriers and facilitators to integrated hypertension-HIV management in Ugandan HIV clinics using the Consolidated Framework for Implementation Research (CFIR)

**DOI:** 10.1101/19013920

**Authors:** Martin Muddu, Andrew K. Tusubira, Brenda Nakirya, Rita Nalwoga, Fred C. Semitala, Ann R. Akiteng, Jeremy I Schwartz, Isaac Ssinabulya

## Abstract

**Background:** Persons Living with HIV (PLHIV) receiving antiretroviral therapy have increased risk of cardiovascular disease (CVD). Integration of services for hypertension (HTN), the primary CVD risk factor, into HIV clinics is recommended in Uganda. Our prior work demonstrated multiple gaps in implementation of integrated HTN care along the HIV treatment cascade. In this study, we sought to explore barriers to, and facilitators of, integrating HTN screening and treatment into HIV clinics in Eastern Uganda.

**Methods:** We conducted a qualitative study at three HIV clinics with low, intermediate, and high HTN care cascade performance, which we classified based on our prior work. Guided by the Consolidated Framework for Implementation Research (CFIR), we conducted semi-structured interviews with health services managers, health care providers and hypertensive PLHIV (n=83). Interviews were transcribed verbatim. Three qualitative researchers used both deductive (CFIR model-driven) and inductive (open coding) methods to develop relevant codes and themes. Ratings were performed to determine valence and strengths of each CFIR construct regarding influencing HTN/HIV integration.

**Results:** Of the 39 CFIR constructs assessed, 17 were relevant to either barriers or facilitators to HTN/HIV integration. Six constructs strongly distinguished performance and were barriers, three of which were in the Inner setting (Organizational Incentives & Rewards, Available Resources, Access to Knowledge & Information); two in Characteristics of individuals (Knowledge & Beliefs about the Intervention and Self-efficacy) and one in Intervention characteristics (Design Quality & Packaging). Four additional constructs were weakly distinguishing and negatively influenced HTN/HIV integration. There were four facilitators for HTN/HIV integration related to the intervention (Relative advantage, Adaptability, Complexity and Compatibility). The remaining four constructs negatively influenced HTN/HIV integration but were non-distinguishing.

**Conclusion:** Using the CFIR, we have shown that while there are modifiable barriers to HTN/HIV integration in the Inner setting, Outer setting, Characteristics of individuals and implementation Process, HTN/HIV integration is of interest to patients, health care providers and managers. Improving access to HTN care among PLHIV will require overcoming barriers and capitalizing on the facilitators using a health system strengthening approach. These findings are a springboard for designing contextually appropriate interventions for HTN/HIV integration in low- and middle-income countries.

**Contribution to the literature:**

- We used the widely used and validated CFIR to assess the HIV program for HTN/HIV integration.
- To our knowledge, this is the first study to explore barriers and facilitators to integrating hypertension screening and treatment into HIV clinics using the CFIR.
- The barriers and facilitators identified are a basis for designing contextualized implementation interventions for HTN/HIV integration in Uganda and other LMIC using a health system strengthening approach.

## Background

Persons living with HIV (PLHIV) and receiving antiretroviral therapy (ART) are at increased risk of cardiovascular disease (CVD) due to ART, HIV infection itself, aging, chronic inflammation, and immune reconstitution [1-3]. In Uganda, approximately 1/3 of PLHIV aged ≥18 years have hypertension (HTN), the leading cause of CVD and preventable mortality [4-12]. PLHIV with HTN have an increased risk of mortality compared to HIV negative persons [13].

The World Health Organization (WHO) and Uganda Ministry of Health (MoH) consolidated guidelines for HIV Care and Treatment recommend that all PLHIV should be screened for HTN at every visit to the HIV clinic. PLHIV who are diagnosed with HTN should receive treatment for both HIV and HTN as integrated services [14, 15]. HTN/HIV integration provides patient-centred care compared with vertical programs and increases efficiency, by eliminating fragmentation and duplication of services [16]. However, there is little empiric evidence describing implementation of this policy in Uganda.

HTN/HIV integration has been attempted in Uganda and Malawi by the SEARCH trial and the Lighthouse Trust program, respectively. However, although HTN screening was achieved for all patients in the HIV clinics under these programs, control of HTN among PLHIV who received antihypertensive medication remained suboptimal at about 30% [5, 17].

We recently conducted a retrospective cohort study of 1649 newly enrolled PLHIV. We mapped the parallel care cascades for HTN and HIV within three high volume HIV clinics (average 3600 PLHIV) in Eastern Uganda and demonstrated suboptimal HTN screening, one year retention in HTN care, and HTN control of 27%, 57% and 24% respectively among PLHIV within a successful HIV program that has achieved the two of three UNAIDS 90-90-90 goals [18]. As a follow-up to that study, the present qualitative study sought to determine barriers to, and facilitators of, integrating screening and treatment of HTN into HIV clinics in Eastern Uganda. Understanding the barriers and facilitators would inform the design of contextually appropriate implementation interventions for HTN/HIV integration in Uganda.

## Methods

### Study design

Using data from our previous retrospective cohort study, we graded HTN care cascade performance among the three HIV clinics as high, intermediate and low according to achievement in screening, diagnosis, initiation of treatment, retention, monitoring and control. We conducted key informant interviews (KIIs) with the district health officer (DHO), each health facility manager (n=3), lead nurse at each HIV clinic (n=3), the lead clinicians at Nagongera and Mulanda HIV clinics (n=2), and two clinicians at TASO Tororo. Additionally, to obtain patients’ information about integrated HTN/HIV services, we conducted focus group discussions (FGDs) and in-depth interviews (IDIs) with hypertensive PLHIV at each HIV clinic.

We utilized the Consolidated Framework for Implementation Research (CFIR) to explore barriers to, and facilitators of, HTN/HIV integration [18]. CFIR provides a pragmatic structure for identifying potential influences on implementation of interventions in health systems at multiple levels [19, 20]. CFIR organizes conceptual elements across theories and disciplines into 39 constructs which are then organized in five key domains. All constructs interact to affect the process and effectiveness of implementation [21]. CFIR’s five major domains include: Intervention characteristics, Outer setting, Inner setting, Characteristics of individuals, and Implementation Process [22]. (Figure 1). Barriers to, and facilitators of, HTN/HIV integration (Intervention) were compared across the three HIV clinics to understand the strength of their influence on HTN screening and treatment among PLHIV.

**Figure 1.**
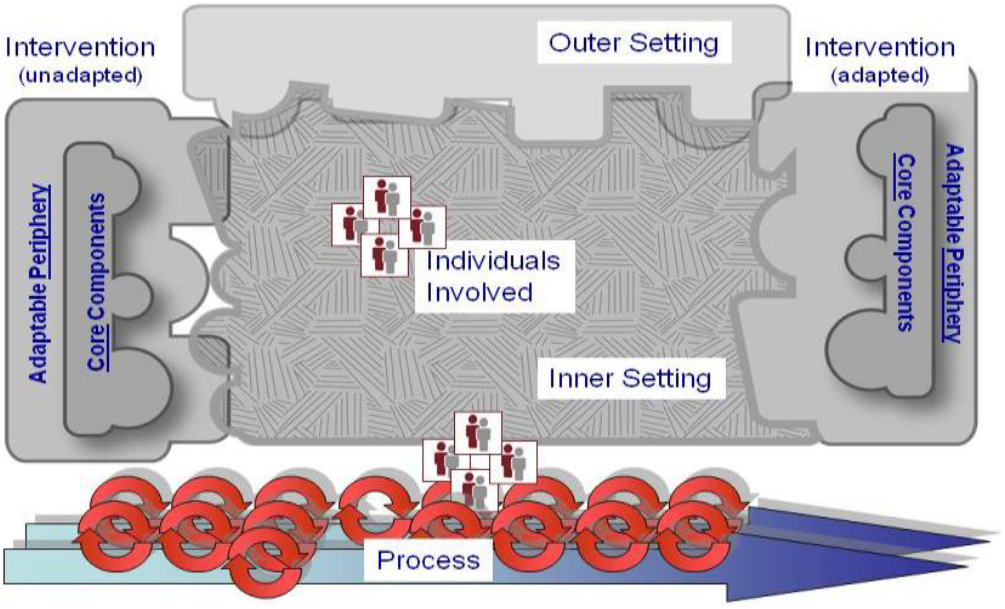
CFIR domains and their relationship. CFIR Wiki.net

### Study setting

We conducted this study at three HIV clinics (The AIDS Support Organization (TASO) Tororo, Nagongera Health Centre IV and Mulanda Health Center IV) and the District Health Office of Tororo District, Eastern Uganda.

The Ugandan public-sector healthcare system is hierarchical in nature and comprises the national (Ministry of Health [MoH]), sub-national (Regional), district, health facility and community levels. MoH develops guidelines for health services and rolls them out through the regions to the districts. The District Health Team (DHT) leads and coordinates guideline implementation at health facilities.

HIV clinics are the designated treatment centers for HIV, HIV-associated opportunistic infections and other HIV-associated co-morbidities. They are physically situated as outpatient departments within health centers and hospitals. We selected these three HIV clinics because they are the largest in Tororo district, providing care to approximately 7500, 1400, and 1100 PLHIV, respectively. They are housed within public health facilities with support from both the Government of Uganda and the President’s Emergency Fund for AIDS Relief (PEPFAR). The clinics are staffed by various cadres of health workers including clinicians, nurses, midwives, and HIV peer counsellors. Each HIV clinic offers a full spectrum of HIV services including screening, ART, viral load testing, as well as screening for and treating opportunistic infections. Each also has the mandate to screen for, and manage NCDs such as HTN and diabetes. Based on existing processes in place at the time of data collection, within a given clinical encounter, BP is measured by the clinician at his/her discretion [18]. If a patient is diagnosed with HTN (by measurement or previous history), the clinician typically prescribes both ART and antihypertensive medicine simultaneously and the client is given one follow-up appointment for both conditions. All medicines at facility pharmacies are obtained from the centralized National Medical Stores (NMS). PEPFAR provides funds to NMS to procure medicines specifically for HIV and opportunistic infections. Medicines for HTN and other NCDs are procured on request by the health facilities via general funds allocated to each health facility by MoH. If medicines are out of stock at the facility pharmacy, the patient is advised to purchase them from a private sector pharmacy of the patient’s choice.

Clinic providers are routinely oriented to national HIV treatment guidelines which recommend screening for NCDs and their risk factors. In addition, clinical support discussions about challenging HIV/NCD cases were also used to build capacity for NCD/HIV integration among clinicians.

### Study participants and Sampling

We interviewed purposively selected healthcare providers and patients living with both HIV and HTN. Eligible healthcare care providers were individuals who had responsibility to treat patients in the HIV clinics or leadership roles at the respective health facilities or at the district health office. These health care providers were knowledgeable about and actively involved in HIV service delivery at their clinics or at the district health office. Eligible patients were PLHIV with hypertension attending one of the three HIV clinics. Patients with a mental disability were excluded.

### Data collection

We used semi-structured interview guides developed based on the five domains of CFIR [19]. The interviews had open ended questions reflecting patient, provider and health care managers’ perspectives and perceptions about HTN/HIV integration. Prior to data collection, we pretested the interview guides with health care providers and hypertensive PLHIV at TASO Tororo who were not participating in the study. We conducted six FGDs (two per HIV clinic) with patients in each group consisting of ten hypertensive PLHIV. We conducted twelve IDIs with hypertensive PLHIV, four at each HIV clinic and eleven KIIs. We used patient IDIs to obtain individual lived experiences, while FGDs explored shared experiences among hypertensive PLHIV.

### Data analysis

#### Qualitative data coding

After transcribing, a research team with expertise in social sciences, public health and clinical care was established comprising three members (AKT, DBN, RN), who conducted thematic content analysis. The team coded transcripts using both deductive (guided by CFIR as a coding framework) and inductive (through open coding to obtain new themes that may arise) approaches. The coding process was guided by the consensual qualitative research (CQR) procedure [23].

First, each research team member read three transcripts independently and identified preliminary codes. Through a series of meetings, discussing coding differences, an initial codebook was agreed upon. To organize and manage the large amount of data, all transcripts were then coded utilizing Atlas.ti (version 7) software while applying the codebook and giving an allowance for new codes. An external researcher independently coded six of the transcripts to establish inter coder reliability (Kappa 0.80). A final codebook and subthemes were resolved by the researchers through more meetings and these were mapped to the CFIR domains and constructs (Table 3).

#### Defining unit of analysis and performance criteria

The three clinics were our units of analysis. We used data from our recent retrospective cohort study [18] which assessed performance of each HIV clinic across HTN care cascade steps including: Screening, Diagnosis, Initiation of Treatment, Retention into care, Monitoring and BP Control (This assessment is also highlighted in figure 2). Generally, the most significant care gaps were identified in Screening and BP control[18]. Basing on the cascade performance reported, we classified the three HIV clinics as high, intermediate, or low performing. (Figure 2).

**Figure 2.**
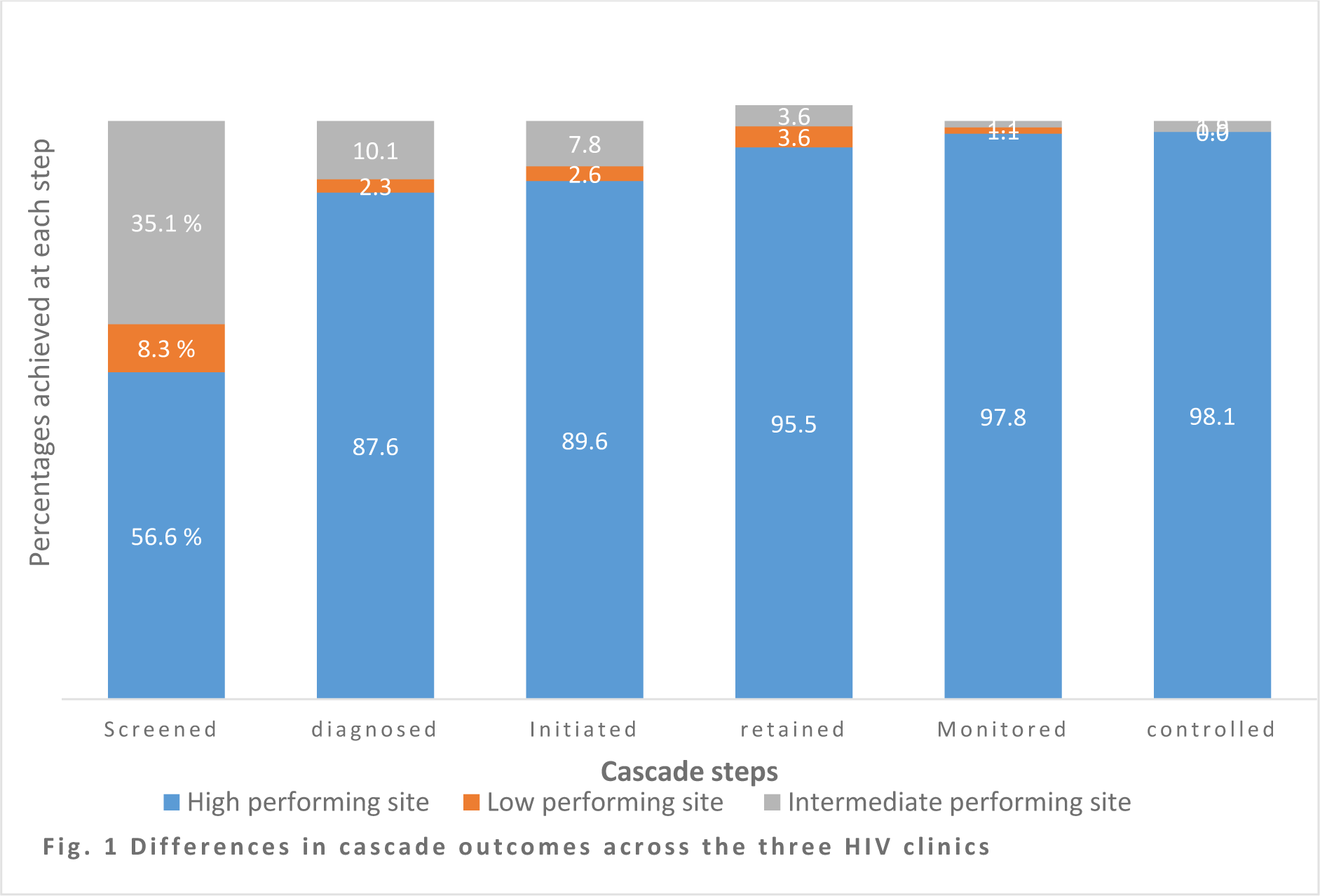
Differences in cascade outcomes across the three HIV clinics included in the study.

#### Rating the CFIR constructs and interpretation

Ratings were performed to determine *valence*, which assesses whether the construct had a positive, neutral, or negative influence on implementation of integrated HTN/HIV care, and *strength* which is the degree of its influence. The coded text was then subjected to a rating process based on criteria shown in Table 1 [24]. We used a consensus process to assign a rating to each construct obtained from each site based on the coded text. We based the rating of constructs on the level of agreement among study participants interviewed, strength of language, and use of concrete examples to emphasize responses (Table 1). Positive influence indicated a facilitator of HTN/HIV integration while negative influence indicated a barrier. After rating all constructs obtained from the three HIV clinics, we developed a matrix that listed the ratings for the CFIR construct for each of the clinics. We then focused our analysis on discerning patterns across the three HIV clinics. Using relative rating, but not absolute figures, we compared ratings across the HIV clinics and determined which constructs distinguished performance, either strongly or weakly, or did not distinguish but influenced performance either negatively or positively. Complexity was ‘reverse rated’ to be consistent with the other constructs; i.e., a positive sign denotes perception of simplicity and a negative sign denotes complexity in implementation [22, 24, 25]. (Table 3). We extracted specific quotations from the transcripts illustrating verbatim expressions of matters that appeared important. We followed the StaRI checklist in developing the manuscript [26].

**Table 1:**
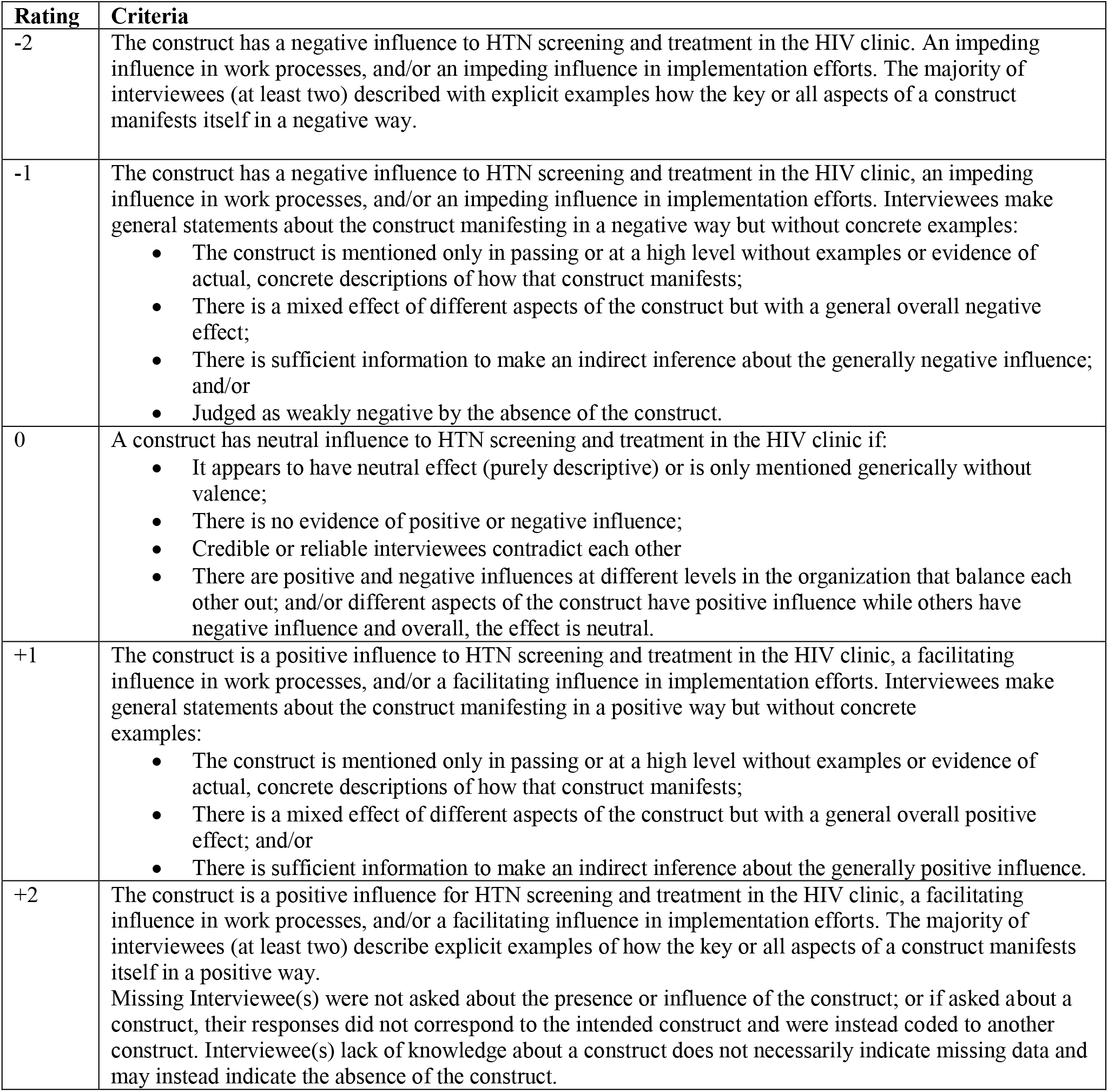
Criteria used to assign ratings to the CFIR constructs that influence screening and treatment of HTN in the HIV clinics.

| Rating | Criteria |
| --- | --- |
| -2 | The construct has a negative influence to HTN screening and treatment in the HIV clinic. An impeding influence in work processes, and/or an impeding influence in implementation efforts. The majority of interviewees (at least two) described with explicit examples how the key or all aspects of a construct manifests itself in a negative way. |
| -1 | The construct has a negative influence to HTN screening and treatment in the HIV clinic, an impeding influence in work processes, and/or an impeding influence in implementation efforts. Interviewees make general statements about the construct manifesting in a negative way but without concrete examples:<br>• The construct is mentioned only in passing or at a high level without examples or evidence of actual, concrete descriptions of how that construct manifests;<br>• There is a mixed effect of different aspects of the construct but with a general overall negative effect;<br>• There is sufficient information to make an indirect inference about the generally negative influence; and/or<br>• Judged as weakly negative by the absence of the construct. |
| 0 | A construct has neutral influence to HTN screening and treatment in the HIV clinic if:<br>• It appears to have neutral effect (purely descriptive) or is only mentioned generically without valence;<br>• There is no evidence of positive or negative influence;<br>• Credible or reliable interviewees contradict each other<br>• There are positive and negative influences at different levels in the organization that balance each other out; and/or different aspects of the construct have positive influence while others have negative influence and overall, the effect is neutral. |
| +1 | The construct is a positive influence to HTN screening and treatment in the HIV clinic, a facilitating influence in work processes, and/or a facilitating influence in implementation efforts. Interviewees make general statements about the construct manifesting in a positive way but without concrete examples:<br>• The construct is mentioned only in passing or at a high level without examples or evidence of actual, concrete descriptions of how that construct manifests;<br>• There is a mixed effect of different aspects of the construct but with a general overall positive effect; and/or<br>• There is sufficient information to make an indirect inference about the generally positive influence. |
| +2 | The construct is a positive influence for HTN screening and treatment in the HIV clinic, a facilitating influence in work processes, and/or a facilitating influence in implementation efforts. The majority of interviewees (at least two) describe explicit examples of how the key or all aspects of a construct manifests itself in a positive way.<br>Missing Interviewee(s) were not asked about the presence or influence of the construct; or if asked about a construct, their responses did not correspond to the intended construct and were instead coded to another construct. Interviewee(s) lack of knowledge about a construct does not necessarily indicate missing data and may instead indicate the absence of the construct. |

**Table 2:** Number of participants involved in this study, by interview type and location.

| **Data collection methods** | **Mulanda health center IV** | **Nagongera health center IV** | **TASO Tororo** | **District Health Team (DHT)** | **Total participants** |
| --- | --- | --- | --- | --- | --- |
| Focus Group Discussion (FGDs) for patients | 20 | 20 | 20 | 0 | 60 |
| In-depth Interviews (IDIs) for patients | 4 | 4 | 4 | 0 | 12 |
| Key Informant Interviews (KIIs) for health care providers | 3 | 3 | 4 | 1 | 11 |
| **Total number of participants** | **27** | **27** | **28** | **1** | **83** |

**Table 3:** Ratings assigned to CFIR construct by study site.

|  | High performing site (C) | Intermediate performing site (B) | Low performing site (A) | Distinguishing constructs |
| --- | --- | --- | --- | --- |
| **I. INTERVENTION CHARACTERISTICS** |  |  |  |  |
| Relative Advantage | +2 | +2 | +2 | Not |
| Adaptability | +2 | +2 | +2 | Not |
| Design Quality & Packaging | +1 | - 2 | -2 | Strongly |
| Complexity | +1 | +1 | +1 | Not |
| **II. OUTER SETTING** |  |  |  |  |
| Patient Needs & Resources | -1 | -2 | -2 | Weakly |
| Peer Pressure | - 2 | -2 | -2 | Not |
| External Policy & Incentives | -1 | -1 | -2 | Weakly |
| **III. INNER SETTING** |  |  |  |  |
| *Implementation Climate* |  |  |  |  |
| Compatibility | + 2 | +2 | +2 | Not |
| Relative Priority | -1 | -2 | -2 | Weakly |
| Organizational Incentives & Rewards | +1 | -2 | -2 | Strongly |
| *Readiness for Implementation* | *Tangible and immediate indicators of organizational commitment to its decision to implement an intervention.* |  |  |  |
| Available Resources | +2 | -2 | -2 | Strongly |
| Access to Knowledge & Information | + 2 | -1 | -1 | Strongly |
| **VI. CHARACTERISTICS OF INDIVIDUALS** |  |  |  |  |
| Knowledge & Beliefs about the Intervention | 1 | 1 | -2 | Strongly |
| Self-efficacy | +1 | -2 | -2 | Strongly |
| **V. PROCESS OF IMPLEMENTATION** |  |  |  |  |
| Executing | -1 | -1 | -2 | Weakly |
| Reflecting & Evaluating | -2 | -2 | -2 | Not |
| Engaging: Opinion leaders | -1 | -1 | -1 | Not |

## Results

TASO Tororo was the highest performing for all the six cascade steps. Mulanda HC IV was intermediate for screening, diagnosis, initiation of treatment, and control but lowest for retention and monitoring. Nagongera HC IV was the lowest for all the six cascade steps having achieved the same with Mulanda HC IV in retention and monitoring. (Figure 2).

Apart from the DHT member, the male to female ratios of participants were 1:1 in all FGDs, IDIs and KIIs. (Table 2).

Of the 39 CFIR constructs assessed, 17 were relevant to either barriers or facilitators to HTN/HIV integration. We found six constructs which strongly distinguished performance, three of which were in the Inner setting (Organizational Incentives & Rewards, Available Resources, and Access to Knowledge & Information); two in Characteristics of individuals (Knowledge & Beliefs about the Intervention and Self-efficacy) and one in Intervention characteristics (Design Quality & Packaging). Four additional constructs distinguished performance weakly. All ten distinguishing constructs negatively influenced HTN/HIV integration. There were four constructs [Relative advantage, Adaptability, the simplicity (non-complex nature) of the intervention and Compatibility] which had positive influence for HTN/HIV integration at all the three HIV clinics but did not distinguish performance. These four factors related to the intervention are the key facilitators for HTN/HIV integration. The remaining three constructs negatively influenced HTN/HIV integration but were non-distinguishing. (Table 3). Below, we present the detailed results in the context of the five CFIR domains.

### Intervention Characteristics

***Relative advantage*** was not a distinguishing construct. Healthcare providers at all three HIV clinics perceived HTN/HIV integration as a *relative advantage* and more effective compared to alternative modes of care for hypertensive PLHIV. Health care providers noted that integrated HTN/HIV was less time consuming since patients received care for both HTN and HIV in the same clinic on the same appointment date, hence reducing costs on transport and improving retention.

*“It is extremely important because if I were a patient having two chronic conditions, I would not want to spend my time going to a hospital for condition A and then go to another for condition B. HIV clinics should be a one-stop centre for our patients so that when they come for ART, they get other drugs at one point so it minimizes the movement. It is extremely important to integrate; it helps to reduce the loss to turn up because you give the patient same appointment. The appointment they are supposed to come for their ARVs is the same day they are supposed to come for their check-up and refills of hypertension medicines: this saves time on the side of clients*.*”* (KII, health facility manager HIV clinic C)

Similarly, ***adaptability*** was not a distinguishing construct since respondents at all the clinics perceived that HTN/HIV integration fits within their routine care provision. This was another facilitator for HTN/HIV integration. Health care providers stated that: HTN services can be tailored and refined to meet health needs of PLHIV. Additionally, HIV clinics have the potential and capacity to implement HTN/HIV integrated services:

*“With availability of resources, integration of hypertension care in our HIV clinics fits and is appropriate for the patients with both conditions*.*”* (KII, Member of the DHT).

*“Yes, hypertension management fits very well within our HIV care programs. This is because to manage hypertension you need trained staff, adequate supplies and space. … and I am happy to inform you that we have three doctors in this small facility*.” (KII, health facility manager HIV clinic C).

Inadequacies in the ***design quality and packaging*** of HTN/HIV integration especially when the program was being initiated was a barrier to the implementation. Health care providers reported that the implementation of HTN/HIV integration policy by MoH was suboptimal. This strongly distinguishing construct mainly affected the low performing clinics. Health care providers at these clinics reported insufficiencies in the preparation, packaging and support for HTN/HIV integration including lack of: staff and professional training systems such as initial orientation to the new health guidelines; clear mechanisms on how resources will be distributed at these facilities; reliable supplies; and necessary materials to implementing the program. These healthcare providers noted that:

*“The introduction of this program was not well communicated to the staff. We only got to know that we are supposed to provide HTN services but not oriented …, not even a workshop. A clear plan for HTN service provision should have been given to us and may be an official launch involving patients and we healthcare providers. … so, I think orientation on the management of hypertension within the clinics is important but we have not had it*.” (KII, facility manager, HIV clinic A).

At the intermediate site it was noted:

*“Since we receive supplies from NMS as one batch, we have a challenge on how some of the supplies needed for HTN management should be distributed at Outpatient Department (OPD) and also at the HIV clinic”* (KII, facility manager, HIV clinic B).

Although the presentation and design of the integration were cited as insufficient, healthcare providers at the high implementing HIV clinic noted that they receive some support through their organisation arrangements which has helped them to get acquainted with the HTN/HIV integration program.

*“We didn’t have any capacity building at the start. However, as an organisation, we regularly have Continuous Medical Education (CME) on non-communicable diseases to boost the staffs’ knowledge. So we got some information on the suggested HTN/HIV integration strategies*.*”* (KII, clinician, site C).

***Complexity*** was not a distinguishing construct perceived to facilitate HTN/HIV integration. Across the HIV clinics, healthcare providers perceived provision of HTN care services as a task which was not complex and that activities for integrated HTN/HIV care were straightforward. However, an increased number of hypertensive PLHIV at the HIV clinics with few healthcare providers would increase provider work load and complexity of handling all patients. A member of the DHT stated:

*“For now, running the HIV clinics when providing care for opportunistic infections (OIs) and other NCDs isn’t difficult at these clinics. It has been done before. We have had HIV implementing partners who support say TB care for patients with HIV. Although …. when patients are many, it could be challenging for the few staff to offer care for the many tasks*.*”*(KII, DHT).

### Outer Setting

***Patient needs and resources*** was a barrier to HTN/HIV integration and weakly distinguished performance across the three HIV clinics. Although HTN/HIV integration was noted as important, both health care providers and patients were in agreement that, to a large extent, HTN services at the HIV clinics were suboptimal to meeting the needs of PLHIV. Providers at lower performing HIV clinics described HTN services as the least prioritized and that they were not well-equipped to meet the needs of HTN patients and there were no indications of efforts to deal with this barrier. In addition, most patients were not aware of HTN/HIV integration which resulted into low demand for the service.

*“Patients are mainly interested in getting the HIV medicine* [ART] *refills. They do not demand for extra care unless their health has deteriorated. Most are not aware of other clinical service we provide including HTN management*.*”* (Lead nurse HIV clinic B).

*“We also like to work on all patients’ conditions but we are sometimes limited by resources. Even now, not much is being done to support HTN integration*.*” (KII, DHO)*.

Besides, several patients interviewed at all the clinics mentioned having little knowledge on HTN/HIV integration and were seeking HTN care at other health facilities.

*“Save for the good HIV care, it [HTN management] is almost not provided at this centre. It was once when the doctor told me to take some tablets for hypertension. … yet when we go for hypertension treatment at other health facilities, they always check your blood [pressure] and even tell you how you are doing. So, I seek treatment outside this centre*.*” (IDI 1, HIV clinic A)*.

Because the low implementing HIV clinics were not in a position to meet PLHIV’s needs for HTN management, hypertensive PLHIV were often referred to other health facilities, a strategy which HTN patients were not comfortable with.

*“I was sent to Tororo general clinic (private for profit) to get checked and I was diagnosed with hypertension. I have the results with me but I’m always told to go back for care at the hospital*.*” (P2, FGD 2, HIV clinic A)*.

***Peer pressure*** was not a distinguishing construct and presented as a negative influencer to HTN/HIV integration. Health care providers were not aware of any HIV clinics implementing HTN/HIV integration to learn or derive competitive pressure from:

*“I have not heard of any facility within or around the district [that is integrating HTN care]. Probably they are doing so, but there is no model health facility I can mention that we can learn from nor healthcare provider to share their experiences and best practices for this program as per now*.*” (KII*, lead clinician HIV clinic C).

**External Policy and Incentives** was a weekly distinguishing construct that negatively influenced HTN/HIV integration. There were few external strategies for HTN/HIV integration through policies and guidelines. Although at the high performing site, healthcare providers stated implementing the national HIV guidelines, their emphasis was on the HIV component:

*“This site … offers a comprehensive HIV care program with focus both on prevention, care and treatment. Currently we are implementing the 2016 National guidelines for HIV/AIDS with emphasis on test and treat*.*”* (Lead clinician HIV clinic C).

This construct was highlighted by healthcare providers at the low performing site that they lacked comprehensive guidelines for HTN/HIV integration:

*“We also lack specific standard operating procedures or documents to be followed in the HTN/HIV integration*.*”* (KII, Lead clinician HIV clinic A).

Besides, healthcare providers also reported insufficient support supervision from the health authorities including MoH, DHT, and implementing partners in relation to HTN management. Relatedly, there were no opportunities to review and support progress of HTN services at the HIV clinics apart from HIV care and supplies.

*“No! We have not received any supervision at the HIV clinic, may be at the OPD [outpatient department]. At least for the past seven months I have been here, I have not seen any. Besides, when we are asked about the clinic, we are often asked about HIV care and supplies”. (KII*, Lead nurse HIV clinic A).

### Inner setting

We identified three sub-constructs under *Implementation Climate* that were relevant to HTN/HIV integration. These were: *compatibility, relative priority and organization incentives and rewards*. **Compatibility** was not a distinguishing construct but a potential facilitator. Healthcare providers perceived that HTN/HIV integration was compatible and would fit within the existing workflows at the HIV clinics. One of the healthcare providers noted:

*“Yes, hypertension services do fit within our routine HIV care service provision. Our work within the HIV clinic is systematic and even hypertension services can be incorporated. We sometimes have BP measured at triage as we measure the patient weight and mid-upper arm circumference”*. (KII, lead clinician HIV clinic A).

**Relative priority**, was a weakly distinguishing construct which negatively influenced HTN/HIV integration. Although health care providers reported that HTN management would fit within the HIV clinic workflow, we found a *relatively lower priority* attached to HTN management by the healthcare providers compared to HIV care and management of opportunistic infections. Healthcare providers reported irregular provision of HTN services at the HIV clinic and providers from the low and intermediate performing facilities explicitly stated that:

*“Basically, we provide the necessary HIV care services and also include care for opportunistic infections like TB and other STIs. Of course, one big challenge is that these other added services may not be frequently provided as expected. and often patients are many; much work to do, so we prioritize HIV care”*. (KII, lead clinician HIV clinic B).

*“We often check the blood pressure for patients with known hypertension and prescribe for them the medicines. However, those ones who are not yet known, we may check once in six months or when they complain with signs and symptoms suggestive of hypertension*.*”* (KII, Lead clinician HIV clinic A).

**Organization incentives and rewards** was a strongly distinguishing construct that negatively influenced HTN/HIV integration. While healthcare providers at the high performing HIV clinic seemed to be motivated by the availability of equipment and other supplies to manage HTN, healthcare providers at the lower preforming sites expressed the need to receive incentives like functional BP equipment and HTN treatment supplies or rewards to motivate their action for additional HTN services, which in this case were not available:

*“There is no special facilitation to do that extra work. …. when such an intervention is to run, at least an allowance for the added work. Even the medicines and other equipment should be available. But we are still struggling to get better BP machines*.” (KII, Lead clinician HIV clinic A).

*“The HIV clinic does not receive special facilitation* [payment] *for managing hypertension cases*.*”* (KII, Lead clinician HIV clinic B).

The two sub-constructs ***Available resources and Access to knowledge and information*** under the ***Readiness for implementation*** construct were potential barriers to the HTN/HIV integration. These two sub-constructs were strongly distinguishing. While they positively influenced performance at the high performing site, they negatively affected performance at the lower performing sites.

**Available resources** for HTN services within the HIV clinics was a strongly distinguishing factor because the high performing site reported having adequate resources including staff who manage HTN, functional BP machines at each department, and medicines for HTN:

*“Yes, I think we have adequate support. We have resources like drugs, equipment they are there, human resource. I can’t say that we are enough but we can’t fail to manage, however few we are we can’t fail to let a case go, at least we manage the fact that the drugs are there the equipment are there, we don’t have any excuse for not managing a hypertensive person*.*” (KII*, Lead clinician HIV clinic C).

On the contrary, low and intermediate performing HIV clinics reported lack of enough equipment especially functional BP machines. They also experience frequent stockouts of the medicines for HTN and the lack of specific funding towards HTN services:

*“… sometimes the* [BP] *machine at the clinic breaks down and it takes a longer process to get another. Yes. Most times we experience stockout of these HTN drugs, so we end up referring the patients*.*”* (KII, facility manager HIV clinic B).

Similarly, **access to information and knowledge** about HTN/HIV integration strongly distinguished between high and low performing HIV clinics. At the high performing site, healthcare providers reported often having trainings including continuing medical education (CMEs) organised at their facility. Some of the CMEs would be refresher trainings and education on NCDs care including HTN.

*“We always have CMEs on hypertension cases, we have had workshops and trainings, management of OIs and hypertension is part of it, that one has been done, so as far as capacity development is concerned, that has been taken care of*.*” (*KII, Lead clinician HIV clinic C).

On the other hand, healthcare providers at low and intermediate performing sites stated low *access to information and knowledge* about HTN care for PLHIV. They identified lack of trainings, few available trained staff at their sites, and poor learning environment as contributing factors.

*“We have not had [trainings or capacity building sessions on hypertension management] for some time and that is the challenge that we have. You know medicine is dynamic and things keep on changing, management of hypertension could have changed like for other conditions. So I think refresher trainings on management of hypertension is important but we have not had them*.*” (*KII, facility manager A).

Another key informant from the district health office stated:

*“It has been about one to two years back, when some health workers were trained on the management of non-communicable diseases. The training happened, but only targeted clinicians. Truly for us to be effective, we need all the others to be trained. Nurses need the training too*.*” (KII, DHO)*.

### Characteristics of individuals

**Knowledge and beliefs** about HTN/HIV integration was a strongly distinguishing construct and a barrier. At the high and intermediate sites, healthcare providers were acquainted with knowledge and skill to offer HTN services like BP measurement and prescription of medicine while at the low performance site, some of the health care providers lacked the skills to appropriately screen and treat HTN:

*“I realized that health workers would report inconsistent BP measurements from patients. Because if patient comes with results from triage and then I measure again, some results would be different. In case the triage says the blood pressure is high, I would measure again in the clinical room. Okay, there are some acceptable variations but there are those that are out of range. So it creates the need for more trainings*.*”* (KII, lead clinician HIV clinic A).

**Self-efficacy**, a strongly distinguishing construct, was a barrier to HTN/HIV integration at the low and intermediate sites. While healthcare providers at the high performing site expressed confidence in their own ability to screen and treat HTN, some healthcare providers at the lower performing HIV clinics expressed low confidence in their own capabilities to screen and prescribe medicines for HTN. One health care provider stated:

*“Patients often present to us [with] different symptoms, in case you follow only these symptoms, you may think its pressure yet it’s not. I have sometimes used the BP machine, but because I don’t use it frequently, I don’t think I get exact measurements to conclude that one has pressure*.*”* (KII Lead nurse HIV clinic B).

Additionally, clinicians at the low performing site expressed concern that lower cadre healthcare providers and the HIV peer counsellors, who often triage patients, lacked enough expertise and confidence to accurately use BP machines for screening and monitoring patients’ BP.

“… *I was noticing variations in the BP measurements, so I withdrew the machines. I told you we no longer screen at triage, so basically integration of HTN care is not at its best because now we depend on the signs and symptoms, not from the routine screening until we get knowledgeable staff that we can place there [to triage] and they give us the correct BP measurements*.*”* (KII, lead clinician HIV clinic A).

### Implementation process

**Execution** of HTN/HIN integration was a weakly distinguishing construct as responses across the three clinics expressed sub optimal HTN services. Several key informants mentioned there are no documents to guide the implementation of HTN/HIV integration at the HIV clinics. HTN care activities at the HIV clinics appeared to be optional and were reported to remain at the discretion of the healthcare provider on duty as mentioned by healthcare providers at the low performing HIV clinic:

“*… we concentrate on the HIV care package, including some STIs or opportunistic infections. We sometimes include hypertension, but not frequently. Like at triage, screening for weight is a must but for BP measurement, it can be once or twice a month*.” (KII, lead clinician HIV clinic A).

**Reflection and Evaluation**, was not a distinguishing construct but negatively influenced HTN/HIV integration. Healthcare providers reported that there was no formalized monitoring or evaluation process of HTN/HIV integrated services at the three HIV clinic. Besides, there were neither known set targets nor reporting/ feedback mechanisms and tools for HTN/HIV integration:

*“We rarely report on HTN care at HIV clinic. Since the HIV card has no provision for recording HTN data, we record on a separate paper which is added to the patient’s file*.*”* (KII, Lead nurse clinic B).

Another healthcare provider noted that:

*“We need to have HTN evaluation forms at these HIV clinics which we don’t do. For example, medical form 5, are often used up and out of stock, they are almost history at some facilities. Patients are encouraged to buy books …. We don’t have good record system to facilitate quick evaluation*.” (KII, DHO).

Some healthcare providers were concerned that quality improvement for HTN/HIV integration was not done for this program to be fully developed at their HIV clinics.

*“*… *we have not had any specific evaluation for this integration since it was communicated to us. Apart from the usual clinic reports that I send as a clinician, and I may inform them that we have a given number of hypertensive patients… but no specific reports made similar to the ones we make, say for TB*.*”* (KII, lead clinician HIV clinic A).

Under the **Engaging** construct, the **Opinion Leaders sub-construct was** non-distinguishing but negatively influenced HTN/HIV integration across the three sites. Key leaders at health facilities, district level and in some HIV implementing partners were not exhibiting commitment nor involvement in HTN/HIV integration. Besides, none of the HIV clinics had enthusiastic healthcare providers who were committed to coordinating or overseeing the implementation of HTN services.

*“We recently rolled out consolidated HIV management guidelines 2016, laid out how you can manage an HIV patient and they have also put emphasis on non-communicable diseases. …, the main issue is who is following up the implementation of care for non-communicable disease at these HIV clinics? This is because each health program and implementing partner concentrate on their targeted disease outcomes. So, you find that they can under look this integration of HTN care*.*”* (KII, DHO).

## Discussion

This study sought to evaluate factors that influence the integration of screening and treatment of HTN into the HIV program in Uganda, using the CFIR. We used valance rating to identify factors which distinguished performance for integrated HTN/HIV between the high and low performing HIV clinics. We found ten CFIR constructs which distinguished performance, four of which were in the inner setting domain. Six of the constructs distinguished performance strongly while the remaining four weakly. All ten distinguishing constructs negatively influenced HTN/HIV integration at the low and intermediate performing HIV clinics as compared to the high performing clinic.

There were four constructs which positively influenced HTN/HIV integration at all the three HIV clinics but did not distinguish performance. These included: Relative advantage, Adaptability, Complexity of the intervention, and Compatibility of HTN care with existing HIV services. We view these as the key facilitators for integrated HTN/HIV services in our setting. In agreement with the facilitators we identified for integrated HTN/HIV services, there is increasing demand for integrated rather than vertically oriented HTN and HIV services in low-and middle income countries (LMIC) [5, 16, 17, 27-29]. Although some studies have assessed barriers and facilitators for HTN care in the HIV program, this is the first study in Africa to assess factors that influence HTN/HIV integration using the CFIR.

From the perspectives of both patients and providers across all the three clinics, integrated HTN/HIV care is seen to have a relative advantage as compared to vertically oriented programs. Providers agreed that having HTN/HIV integrated services will allow for improved patient-centred care. For example, patients would receive both HTN and HIV services instead of being referred to two different clinics. Furthermore, integration reduces duplication of services, is cost effective and efficient [5, 16, 29]. Our recent work also showed that the HIV cascade results were similar between patients with HIV alone and those with HIV and HTN who received integrated care [18]. Such evidence bolsters the demand for integrated HTN/HIV services from health care providers and patients that we see in the current study. Leveraging HIV programs for HTN care may even provide a spillover effect to the non-HIV population by increasing access to screening, treatment and control of HTN [5, 27].

Adaptability was a positive influencer of integrated HTN/HIV services. Health care providers and leaders, especially in the high performing clinic, perceive that integrated HTN/HIV care can be adapted and tailored to fit the workflow of the HIV clinics to meet patient needs. HIV services were originally established as vertically oriented entities to address the emergency nature of HIV. Now that HIV has evolved into a chronic disease, largely due to these intensive, vertical efforts, leveraging successful HIV programs for NCD integration is highly recommended [16, 29]. A large clinical trial in Uganda that integrated HTN care into the HIV program showed that HTN control is better achieved in the HIV program as compared to the non-HIV population [30]. This is a true demonstration of the adaptability of integrated HTN/HIV service to the HIV program [30].

Most health care providers at all HIV clinics found HTN/HIV integration not to be complex, making it a facilitator. However, providers anticipated that as more dually affected patients are identified, the workload may become overwhelming if not met with a concomitant increase in staffing or resources. Task shifting could assist in mitigating the impact on providers [18]. In parallel with task shifting, as was and is done in the HIV context, programs must continue to strengthen the capacity of the existing clinical workforce to provide integrated care through re-education efforts, continuing medical education (CME), simplified evidence based treatment protocols/algorithms, implementation guidelines and physical and/or digital decision aides [5, 16]. Since Access to information and knowledge was a barrier to HTN/HIV integration, capacity building for HTN and other NCDs in HIV should not only target clinicians but all cadres of staff so that the HTN/HIV services are supported by interdisciplinary teams for sustainability and efficient task shifting [5, 16].

Health care providers noted that HTN/HIV integration was compatible with the existing services in the HIV clinics and workflows. A project in Malawi that integrated screening and treatment of HTN into the HIV clinics demonstrated that HTN care was compatible with HIV services [5]. Indeed, as we demonstrated previously. the care cascades for both conditions can be perfectly aligned [18].

All ten constructs which distinguished performance had a negative influence upon HTN/HIV integration. Design quality and packaging was a barrier to HTN/HIV integration. Health care providers from the lower performing clinics were not oriented on HTN/HIV integration, unlike those at the high performing site. Additionally, Ugandan HIV programs will need to adopt the WHO target of 50% for HTN screening, treatment, retention in care and control. Achieving this will require quality improvement efforts and integration of CVD indicators into routinely collected data at National, regional, district and health facilities [27].

Patient needs and resources weakly distinguished performance for HTN integration. Key resources that hindered performance at the lower performing clinics included BP machines, access to medicines for HTN, and information on HTN/HIV integration. Many patients were not aware of HTN/HIV integration at the HIV clinic and, therefore, could not advocate for the service. Fixed dose combinations of HTN medicines will reduce the pill burden of treating both HTN and HIV. Additionally, differentiated service delivery models and Chronic Care Models (CCM) for integrated HTN/HIV will strengthen treatment adherence and promote retention in care and patient centeredness [14, 15, 17, 29, 31].

Relative priority was a weak barrier to HTN/HIV integration. Health care providers mentioned that HTN screening and treatment among PLHIV are not prioritized. Providers mentioned that BP measurement at HIV clinic is only done for clients who are already confirmed to have HTN or those with symptoms but not all PLHIV. This non-prioritization of HTN services by providers leaves out many patients undiagnosed with HTN since majority have no symptoms especially in the early stages. This approach risks diagnosing patients very late with overt complications of HTN yet PLHIV are already in close contact with the health care system and present an opportunity for CVD screening, treatment and control [29]. Due to suboptimal screening and treatment of HTN among PLHIV in Uganda, awareness of HTN and control remain below 20% [7-12, 18].

Providers at the high performing clinic were motivated by incentives and available resources including functional BP machines, access to medicines for HTN and human resources unlike their counterparts at lower performing clinics who noted that they would derive motivation from the same incentives and resources. Our findings are in agreement with a recent mixed methods study in Nigeria that assessed the capability, opportunity and motivation for HTN/HIV integration found that physical opportunity in form of BP machines and medicines was suboptimal in most HIV clinics [28]. Strategies to improve access to medicines will require prioritization of availability and affordability of standardized selected core medications for HTN [5, 31].

Self-efficacy of health care providers to screen and treat HTN was low in the lower performing HIV clinics unlike in the high performing clinic. Additionally, clinicians at the lower performing clinics expressed lack of confidence in lower cadres of health care providers as far as screening for HTN was concerned. These findings are supported by prior studies that have found low levels of confidence and self-efficacy regarding HTN screening among multiple cadres of health workers [28, 32]. Strategies to address providers’ confidence will rely on improving knowledge and skills through cadre-appropriate training, mentorship and education [5, 16].

Execution of HTN screening and treatment is generally suboptimal since it is at the discretion of the clinician especially at the lower performing HIV clinics. Suboptimal screening leads to low levels of awareness, treatment, retention and control of HTN among PLHIV [7-12, 18]. Thus, there is a need to routinely provide HTN screening and treatment at HIV clinics, through use of standardized evidence based treatment protocols, improved access to medicines, mentorship, target setting, improved systems for monitoring and evaluation, empowering patients, task shifting, differentiated service delivery and community engagement [5, 31].

Participants in our study reported gaps in clinician documentation because providers record clinical data in patients’ personal books. These records leave the clinic with the patient following their visits. Enhanced systems for monitoring and evaluation for HTN in the HIV clinics are critically needed. This gap limits access to patient data and information for programmatic planning and continuous quality improvement. To close this gap, HTN care indicators should be integrated into the electronic medical records (EMR) system that is available and functional at HIV clinics [33].

We acknowledge the paucity of contributions to the present analysis by the many patient participants interviewed for this study. Future implementation science research and analyses should place more emphasis on patients’ perspectives and perceptions on HTN/HIV integration.

### Conclusions

Using the CFIR framework, we have shown that there are modifiable barriers to integrating HTN services into the HIV clinic in the inner setting, outer setting, characteristics of individuals and implementation process. Integrated HTN and HIV care is of great interest to both patients and health care providers. Improving access to HTN and other CVD care among PLHIV will require overcoming these barriers and capitalizing on the facilitators identified using a health system strengthening approach [5]. Findings from this study provide a springboard for designing contextually appropriate multicomponent interventions for HTN/HIV integration in Uganda and other LMICs. To further build the case for integrated HTN/HIV services, future research should determine the cost, cost effectiveness and treatment outcomes for both HTN and HIV from integrated HTN/HIV services [16, 27].

## Data Availability

The datasets used and/or analyzed during the current study are available from the corresponding author on reasonable request.

## List of abbreviations

CFIR: Consolidated Framework for Implementation Research
HTN: Hypertension
PLHIV: Persons Living with HIV
CVD: Cardiovascular Disease
ART: Antiretroviral Therapy
WHO: World Health Organization
MoH: Ministry of Health
FGD: Focus group discussion
KII: Key Informant Interview
IDI: In-depth interview
DHO: District Health Officer
DHT: District Health Team
LMICs: Low-and Middle Income Countries

## Declarations

### Ethics approval and consent to participate

This study was approved by the TASO Institutional Review Board (IRB) number TASOREC/65/17-UG-REC-009 and the Uganda National Council for Science and Technology (UNCST) number SS4583. Permission (written consent) to access the HIV clinics and medical records was sought from the DHO and health facility in-charges respectively. Written informed consent was sought from each participant after explaining the purpose of the study, benefits, approximate time of interview and an assurance of confidentiality of the study results. Selected individuals were asked if they would like to participate, and if the response was affirmative, the interviewer would give them a consent form to be signed as evidence of acceptance to take part in the study. In case a participant was not willing to be audio recorded for key informant or in-depth interviews, interview notes were taken. Privacy was maintained during data collection, analysis and storage. All identifiable information was removed from final records after data collection to ensure participant anonymity and only the core research team and PIs had access to the de-identified data.

### Consent for publication

Not applicable

### Availability of data and materials

The datasets used and/or analysed during the current study are available from the corresponding author on reasonable request.

### Author contributions

MM, IS, JIS and AKT were responsible for the design of the study and interpretation of data. MM, AKT, BN and RN led data collection and interpretation. AKT, BN and RN performed data analysis. ARA participated in study design and data interpretation. FCS participated in the interpretation of data. All authors participated in writing the initial draft of the manuscript. Martin Muddu and Jeremy and Isaac Ssinabulya participated in writing the final manuscript. All authors read and approved the final manuscript before submission.

## Acknowledgement

We are grateful to the following persons for their invaluable support: The District Health Officer and the District Health Team of Tororo District, the staff of TASO Tororo, Nagongera HC IV and Mulanda HC IV HIV clinics and all the research assistants who participated in data collection for this study.

